# Urban-rural differences in pediatric ATV-related trauma in Canada from 2002-2019: A population-based descriptive study

**DOI:** 10.1101/2025.07.17.25331717

**Authors:** Mattea Heck, Shamsia Sobhan, Robert Balshaw, Jonathan McGavock

**Affiliations:** Children’s Hospital Research Institute of Manitoba; Department of Pediatrics, Rady Faculty of Health Sciences, University of Manitoba; George and Fay Yee Centre for Healthcare Innovation, University of Manitoba; Department of Community Health Sciences, Rady Faculty of Health Sciences, University of Manitoba.

**Author notes:** **Address of Correspondence:** Mattea Heck, MBChB, Department of Pediatrics and Child Health Faculty of Health Sciences, University of Manitoba, 511 JBRC. Children’s Hospital Research Institute of Manitoba. 715 McDermot Ave. Winnipeg, Mb R3E 3P4.

**Keywords:** Inequities, geographic, all-terrain vehicles, concussion, pediatric, trauma, emergency medicine

## Abstract

**Objective:** The aim of this study was to describe differences and trends in ATV-related hospitalizations for urban and rural-dwelling youth in Canada.

**Methods:** We conducted a cross-sectional study using administrative hospital abstract data all patients admitted for an ATV-related injury to hospitals in 9 provinces in Canada between 2002 and 2019. The primary exposure was rural residence, defined by postal code. Rural-urban comparisons were stratified by age group: children (<16 years), adolescents (16-20 years) and adults (>21 years). The primary outcome was the incidence of any hospitalization, secondary outcomes were head injury, fractures, crush injury and spinal cord injury..

**Results:** Among 34,390 patients with complete data, 17% were children younger than 16 yrs and 14% were adolescents 16-20 yrs; 78% of children and 85% of adolescents were male, and 47% lived rurally. The incident rate ratio (IRR) for being hospitalized for an ATV-related injury was 5-fold higher for rural children (5.59; 95% CI: 5.30-5.88) and adolescents (5.16; 95% CI: 4.88, 5.47) compared to urban children and adolescents, respectively. The 5-fold higher IRR was also evident for ATV-related fractures among rural children and adolescents. Adolescents had a particularly higher risk for ATV-related crush injuries (IRR: 10.43; 95% CI: 5.74-18.96) and spinal cord injuries (IRR: 5.21; 95% CI: 3.33-8.15) while children were at higher risk of ATV-related head injuries (IRR: 6.55; 95% CI: 5.76-7.46) compared to urban dwelling youth.

**Conclusions:** In Canada, rural children and adolescents were at a very elevated risk of ATV injuries compared to those living in urban centres.

## INTRODUCTION

Tens of thousands of children and adolescents (referred to herein as youth) are hospitalized for all-terrain vehicle (ATV) related injuries in Canada and the US every year(1–4). To reduce ATV injuries in youth, the Canadian Pediatric Society(5) and the American Academy of Pediatrics (AAP)(6) recommend unique policies aimed at youth living in rural areas. Targeted public health policies aimed at rural-based youth are recommended due to differences in ATV exposure, riding patterns and terrain(6, 7). A better understanding of the distribution of ATV-related hospitalizations for urban and rural-dwelling youth could inform public health policies to address these recommendations.

Surveys of pediatric ATV users reveal that riding patterns and risk-taking behaviors are different between rural and urban youth(8). Some data suggest that injury patterns and severity are also different between rural and urban dwelling patients(9, 10). Unfortunately, these studies are restricted to single-centre medical records and are at risk of selection bias, misrepresenting the true inequities in ATV-related injuries among rural dwelling pediatric populations. A comprehensive population-based estimate of the distribution of ATV-related injuries between rural-urban children and adolescents could overcome this gap in pediatric injury epidemiology and inform public health strategies aimed at preventing ATV-related hospitalizations among children and adolescents.

The current study built on previous work from our group(11) and was designed to describe the magnitude and precision of inequities in hospitalizations for ATV-related injuries between rural and urban-dwelling children and adolescents in Canada. Using administrative hospitalization data for ATV-related injuries over an 18-year period, from 9 provinces in Canada, the main research question guiding this study was: Among children and adolescents in Canada, are rates of hospitalizations for ATV-related injuries different for those living in rural areas, compared to those in urban areas? In addition, we assessed if the magnitude of rural-urban inequities in ATV-related hospitalizations were different in age and gender-based sub-groups of the population.

## METHODS

### Data sources and Population

We used a cross-sectional study design embedded within a large national administrative registry of hospitalization records in Canada(12) to answer the main research questions. We followed Lesko’s framework for designing and reporting descriptive epidemiological studies(13). The data management staff within the Canadian Institute for Health Information provided a database for all hospitalizations for persons 1 to 80 years of age admitted to a hospital for an ATV-related injury between January 1^st^ 2002 and December 31^st^ 2019 within the Discharge Abstract Database (DAD)(14). The DAD database contains diagnostic information on all admissions to acute care hospitals in Canada, primary and secondary International Classification of Diseases 10^th^ (ICD-10A) revision codes and demographic information for all patients admitted to hospitals in 9 provinces in Canada. Hospitalization data for the province of Quebec was not available as it does not participate in the registry and there was limited data from hospitals in the Northern territories (Yukon, Nunavut, Northwestern Territories). The ICD-10CA external cause of injury codes for off-road vehicle or snowmobile injuries, used at admission to hospital are provided in Table S1 of the appendix. Staff at the Canadian Institute for Health Information provided access to a password-protected database with de-identified data for age category, three-digit postal codes, sex, ICD-coded injuries, outcomes and co-variates for the analyses described below. The research team did not have access to the entire database from which data were extracted. We followed STROBE(15) and RECORD(16) reporting guidelines to report the results of this study. Analyses were approved by the health research ethics board at the University of Manitoba (HS:26060 ; H2023;205).

### Exposure and sub-groups of interest

The primary exposure of interest was rural residence determined by individual postal codes available at the time of admission. Patients in the comparison group lived in a residence with a postal code from an urban area, defined as a city with >10,000 residents. Comparisons of outcomes between rural and urban residents were done within three age-specific sub-groups. Patients younger than 16 years at the time of the ATV injury categorized as children; patients 16 to 20 years of age were classified as adolescents; and patients over 21 years of age were considered as adults. The age of less than 16 years was used for a sub-group strata as: (1) 16 years of age is the minimum age to obtain a licence for driving a vehicle in Canada and (2) 16 years of age is also the minimum age suggested for riding an ATV in most Canadian provinces (17). The age of 21 years was used to define adults by CIHI and was used in previous studies by our team due to differences in riding patterns around that age range(11).

### Outcomes of Interest

The main outcome of interest was the incidence of any hospitalization for an ATV-injury per 1000 persons. To calculate provincial incident rates of ATV-related hospitalizations, annual age-specific population estimates were provided for each province by Statistics Canada. Secondary outcomes were injury sub-types including fractures, crush injuries, head injuries and spinal cord injuries. The cause and nature of each patient’s injury are coded according to ICD-10-CA codes, and up to 10 injuries coded per patient were provided by the data stewards (eTable 1 for specific codes). ICD10A codes are frequently used in administrative database studies to classify injuries and provide valid estimates of injury severity(18). Head injury was defined as injury to the brain, skull, scalp, or face. Spinal cord injury was defined as injury to the cervical, thoracic, or lumbar spinal cord, or cauda equina. Crush injury was defined as crush injury to the head, neck, thorax, abdomen, pelvis, upper or lower extremity, or multiple body parts. Fractures were defined as fractures to any bone of the upper or lower extremity.

### Demographic and co-variates of interest

The province of hospital where the patient was admitted, year and month of the injury, and sex for the injured patient were provided with the list of ATV-related injuries for each patient. Data for helmet use, and time spent riding an ATV prior to being hospitalized were not available within the registry.

### Patient and Public Involvement

No members of the public or patient partners were involved in this research project.

### Analyses

The sample size of 34,390 was the number of available patients that met inclusion criteria for the proposed analyses. To answer the main research questions, we performed Poisson regression analyses to determine if incident rates of the outcomes of interest were higher among children and adolescents, living in rural areas compared to those in urban areas. Incident rate ratios (IRR) and 95% confidence intervals (CI) were calculated to estimate statistical differences between rural and urban patients. Sub-group analyses for age and gender-specific differences were conducted after interaction terms in sensitivity analyses suggested age or gender-based differences were present. As less than 5% of data were missing from the available database, we did not impute missing data and no sensitivity analyses were performed. The sample was derived from eligible patients within the DAD database for the years of interest. All data were analyzed using R Version 4.2.3 (2023-03-15).

## RESULTS

A flow chart describing the number of patients excluded and the rationale for the final sample size used for these analyses is provided in eFigure 1 of the appendix. Between 2001 and 2020, CIHI provided data for 36,387 patients who were admitted to hospital for one of the four ATV-related injuries of interest (fractures, head injuries, crush injuries and spinal cord injuries) across the 9 provinces and territories. We excluded data from patients with an ICD-10 code suggesting that they were injured after being struck by an ATV (V86.7).

For the primary analyses, 34,390 patients that were admitted to hospital for ATV injury during 2002-2019 had complete data from 9 provinces; 16.8% of participants were children (under 16 years of age), 13.8% were adolescents (aged 16 to 20 years), 69.4% were adults (>21 years); 82.8% were male and 47.1% lived in a rural area (Table 1). The distribution of sex, province and geographic residence was similar across the three study groups. Annual hospitalization rates were relatively stable from 2002-2019 (eFigure 2). The annual incident rate for ATV-related hospitalizations ranged from 4-9/100,000 for children, to 8-21/100,000 for adolescents and 4-8/100,000 for adults during the study period. The annual incident rate per hundred thousand for ATV-related hospitalizations for children and adolescents were highest in the province of Newfoundland and Labrador (range: 11-32/100,000 for children and 25-60/100,000 for adolescents) and lowest in the province of Ontario (eFigure 3).

**Table 1.**
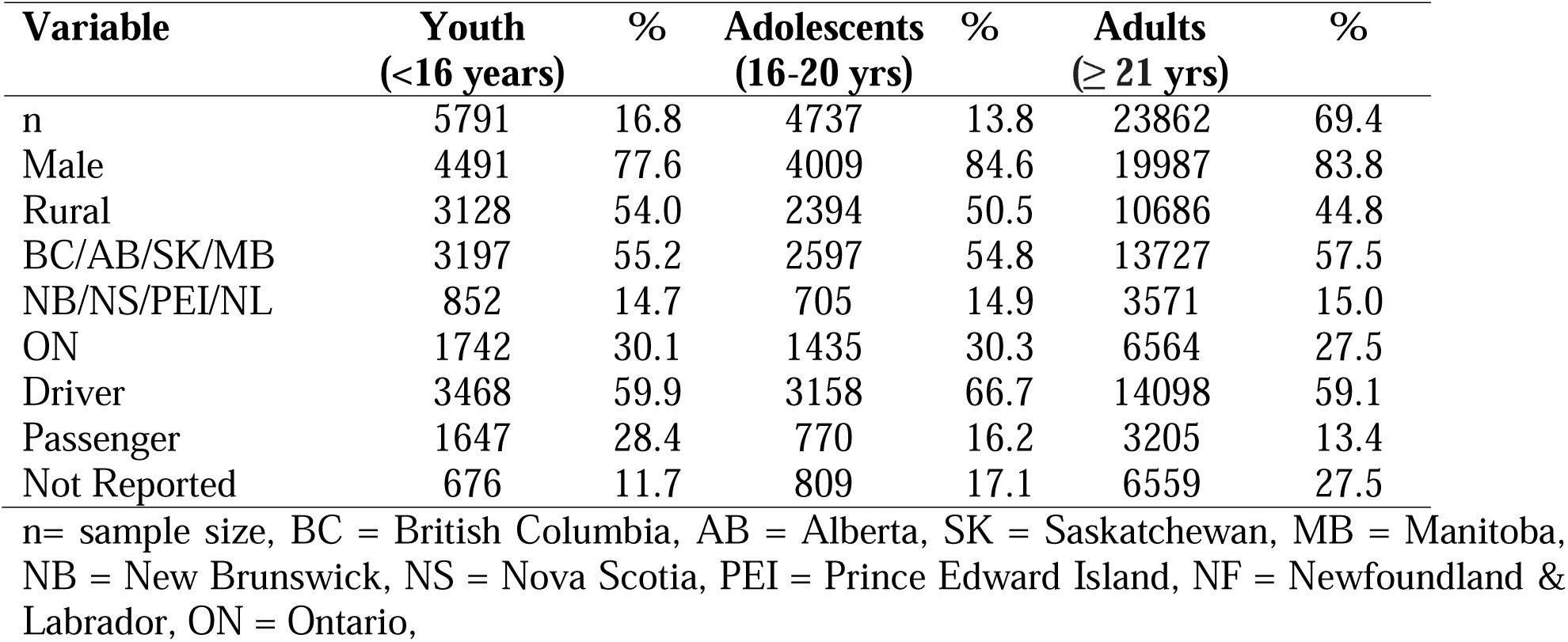
Study participant demographics stratified according to age group.

| **Variable** | **Youth**<br>**(<16 years)** | **%** | **Adolescents**<br>**(16-20 yrs)** | **%** | **Adults**<br>**(≥ 21 yrs)** | **%** |
| --- | --- | --- | --- | --- | --- | --- |
| n | 5791 | 16.8 | 4737 | 13.8 | 23862 | 69.4 |
| Male | 4491 | 77.6 | 4009 | 84.6 | 19987 | 83.8 |
| Rural | 3128 | 54.0 | 2394 | 50.5 | 10686 | 44.8 |
| BC/AB/SK/MB | 3197 | 55.2 | 2597 | 54.8 | 13727 | 57.5 |
| NB/NS/PEI/NL | 852 | 14.7 | 705 | 14.9 | 3571 | 15.0 |
| ON | 1742 | 30.1 | 1435 | 30.3 | 6564 | 27.5 |
| Driver | 3468 | 59.9 | 3158 | 66.7 | 14098 | 59.1 |
| Passenger | 1647 | 28.4 | 770 | 16.2 | 3205 | 13.4 |
| Not Reported | 676 | 11.7 | 809 | 17.1 | 6559 | 27.5 |

n= sample size, BC = British Columbia, AB = Alberta, SK = Saskatchewan, MB = Manitoba, NB = New Brunswick, NS = Nova Scotia, PEI = Prince Edward Island, NF = Newfoundland & Labrador, ON = Ontario,

The distributions of each injury type by age are provided in Table 2. For children and adolescents, the most common ATV-related injuries requiring hospitalization were fractures (children: n=4,745, 81.9%; adolescents: n=3,803, 80.3%) and head injuries (children: n=943, 16.3%; adolescents: n=808, 17.1%). The distribution of injury types were similar across all three age groups. Among the 108 patients under 21 years of age that were hospitalized with a spinal cord injury, rates were 3-fold higher in adolescents, compared to children (0.5% vs 1.6%). The annual percentages change in ATV injuries for rural and urban children and adolescents are shown in eFigure 4, where trends were similar for age and geographic sub groups.

**Table 2.** Injury characteristics for patients admitted to hospital for an ATV injury in Canada.

| **Variable** | **Youth**<br>**(<16**<br>**years)** | **%** | **Adolescents**<br>**(16-20 yrs)** | **%** | **Adults**<br>**(≥ 21 yrs)** | **%** |
| --- | --- | --- | --- | --- | --- | --- |
| n | 5791 |  | 4737 |  | 23862 |  |
| Fractures | 4745 | 81.0 | 3803 | 80.3 | 19493 | 81.7 |
| Crush Injuries | 72 | 1.2 | 49 | 1.0 | 269 | 1.1 |
| Head Injuries | 943 | 16.3 | 808 | 17.1 | 3544 | 14.9 |
| Spinal Cord Injuries | 31 | 0.5 | 77 | 1.6 | 556 | 2.3 |
| Population at risk per<br>year (Range) | 4,782,123<br>(4,697,366-4,988,766) |  | 1,723,455<br>(1,665,165-1,766,779) |  | 19,677,695<br>(17,326,589-22,299,237) |  |

n= sample size

The unadjusted incident rates for rural and urban children and adolescents for ATV-related hospitalizations and specific injuries are presented in Table 3. Geographic and age-specific differences in pediatric ATV-related hospitalizations were relatively similar for all injury types (Figure 1). Compared to urban dwelling children and adolescents, rural dwelling children and adolescents were 5 times more likely to be hospitalized for an ATV injury (IRR: 5.59; 95% CI: 5.30-5.88 and IRR: 5.16; 95% CI: 4.88-5.47 respectively) and experience ATV-related fractures (IRR: 5.48; 95% CI: 5.18-5.80 and IRR: 5.10; 95% CI: 4.78-5.43 respectively) (Table 3). Rural dwelling adolescents were 10-fold more likely to experience a crush-type injury (IRR: 10.43; 95% CI: 5.74-18.96) while rural dwelling children were 6-fold more likely to experience a head injury (IRR: 6.55; 95% CI: 5.76-7.46) compared to urban dwelling adolescents and children respectively. Compared to urban dwelling peers, rates of spinal cord injury were 2-fold (IRR: 2.27; 95% CI 1.07-4.82) and 5-fold (IRR: 5.21; 95% CI: 3.33-8.15) higher for rural-dwelling children and adolescents respectively.

**Figure 1.**
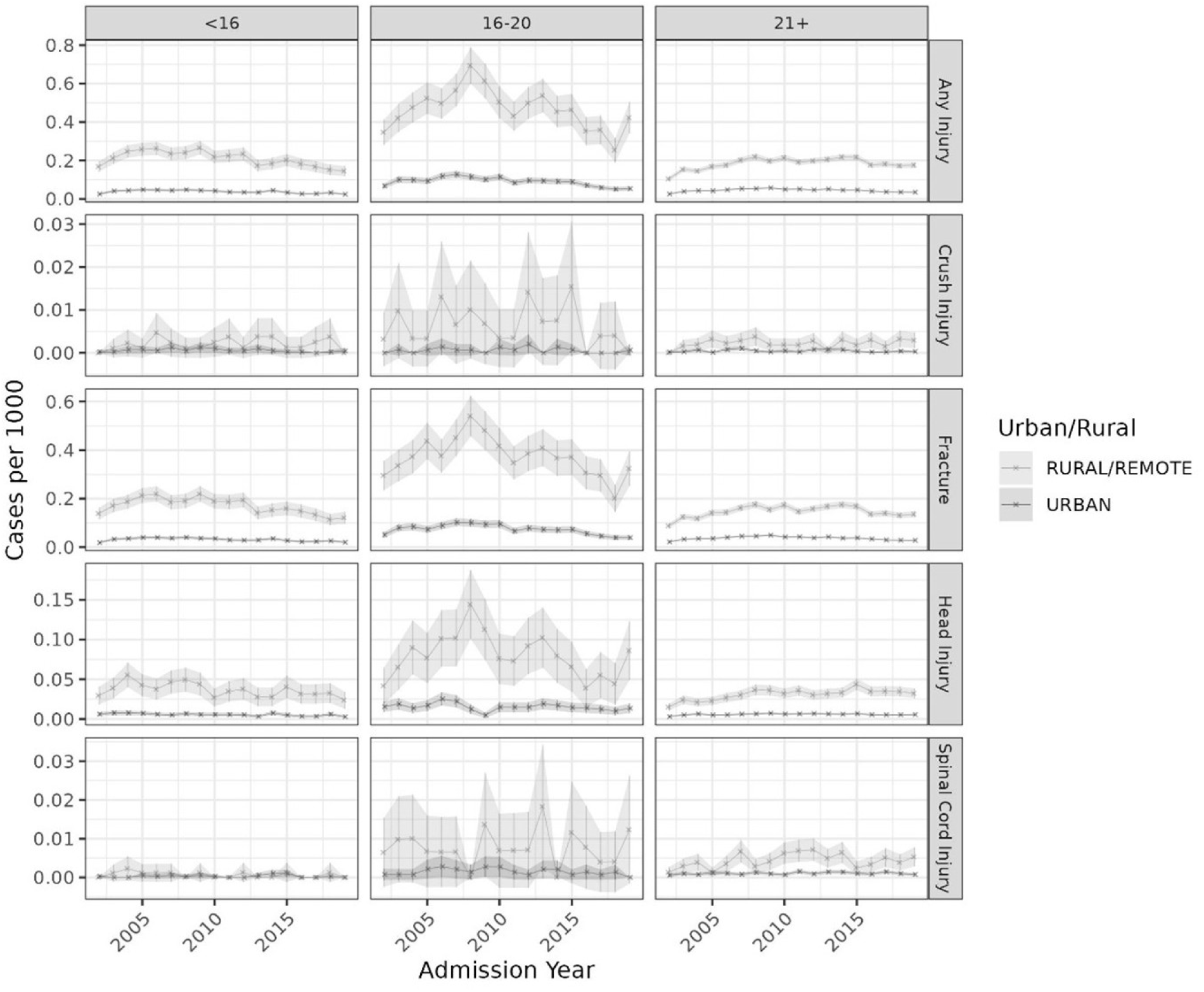
Incident rates for ATV-related injuries and injury types for rural and urban dwelling patients in Canada from 2002-2019, stratified by age group

**Table 3.** Incidence rate per 100,000 and incident rate ratio (IRR) with 95% confidence interval (CI) of rural children and adolescents for hospitalizations for ATV-related injuries.

| Injury Type | Incident Rate per 100,000 |  | IRR (95% CI) |
| --- | --- | --- | --- |
|  | Rural | Urban |  |
| Children (<16 yrs) |  |  |  |
| Total | 21.03 (14.50, 26.56) | 3.74 (2.41, 4.82) | 5.59 (5.30, 5.88) |
| Crush | 0.21 (0.00, 0.47) | 0.06 (0.00, 0.13) | 3.61 (2.27, 5.76) |
| Fracture | 17.08 (11.42, 21.93) | 3.10 (1.84, 4.03) | 5.48 (5.18, 5.80) |
| Head | 3.68 (2.37, 5.54) | 0.56 (0.29, 0.77) | 6.55 (5.76, 7.46) |
| Spinal Cord | 0.07 (0.00, 0.23) | 0.03 (0.00, 0.10) | 2.27 (1.07, 4.82) |
| Adolescents (16-21 yrs) |  |  |  |
| Total | 47.13 (25.37, 69.43) | 9.03 (5.09, 12.75) | 5.16 (4.88, 5.47) |
| Crush | 0.65 (0.00, 1.54) | 0.06 (0.00, 0.20) | 10.43 (5.74, 18.96) |
| Fracture | 37.60 (20.13, 54.00) | 7.30 (3.93, 10.24) | 5.10 (4.78, 5.43) |
| Head | 8.11 (3.87, 14.42) | 1.53 (0.48, 2.53) | 5.27 (4.59, 6.05) |
| Spinal Cord | 0.77 (0.00, 1.83) | 0.15 (0.00, 0.28) | 5.21 (3.33, 8.15) |

Figure 2 illustrates the differences in incident rate ratios for ATV-related hospitalizations for rural dwelling children and adolescents, compared to urban dwelling peers in different provinces. Across all provinces, except New Brunswick, the rates of ATV-related hospitalizations were significantly higher in rural children and adolescents compared to urban peers. The province with the largest rural-urban inequity was Ontario where the incident rate was ∼10-fold higher and lowest in Newfoundland where the incident rate was ∼2.5 fold higher in rural children and adolescents, compared to urban peers. The annual percentage change in ATV-related hospitalizations stratified by rural-urban residence and age (children and adolescents) are provided in eFigure 4 and suggested that rates declined slightly in recent years, in all sub-groups.

**Figure 2.**
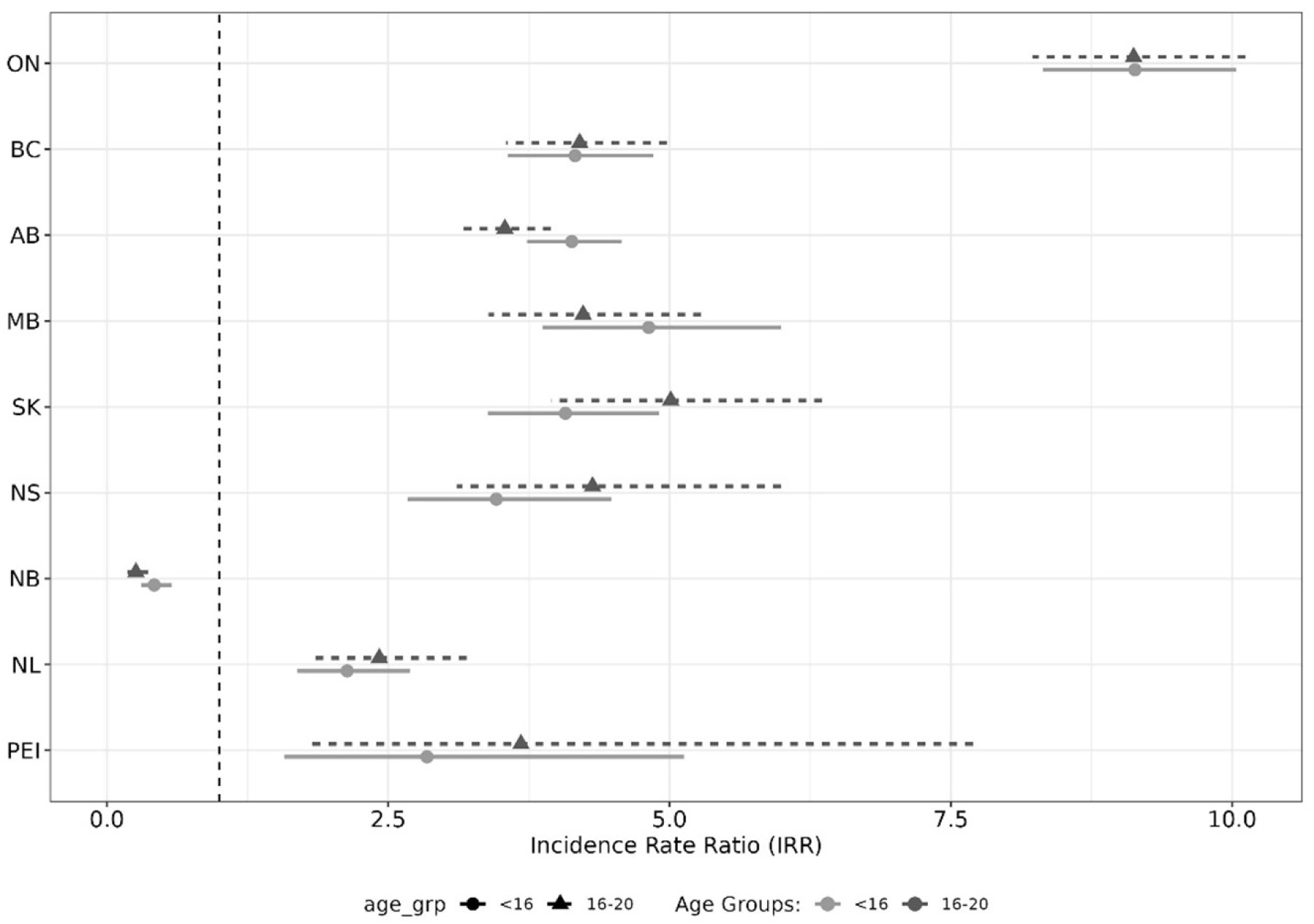
Incident rate ratios for differences in ATV-related hospitalizations for rural compared to urban-dwelling children and adolescents per province.

A summary table for absolute incident rates of ATV-related hospitalizations for boys and girls across all age groups is presented in the appendix (eTable 2). The annual incident rate of being hospitalized for an ATV-related injury was 3-fold higher for male children (range: 6-13/100,000) and 5-fold higher for male adolescents (range: 13-35/100,000) compared to female children (range: 2-4/100,000) and female adolescents (range: 3-7/100,000) respectively. Interaction terms for sex and rural residence suggested that gender differences in ATV-related hospitalization were greater in rural areas compared to urban areas. Specifically, the incidence rate for rural boys was 8-fold higher for children (IRR: 8.38; 95% CI: 7.93-8.85) and adolescents (IRR: 8.36; 7.88-8.88), compared to girls in each age group.

## DISCUSSION

Several novel findings emerged from this descriptive epidemiologic study of ATV-related hospitalizations in 9 Canadian provinces. First, rates of ATV-related hospitalizations, regardless of injury type, are 3-10 times higher among children and adolescents living in rural areas, compared to children and adolescents in urban areas. Second, adolescents aged 16-21 years, display the highest rates of ATV-related hospitalizations, and these rates are highest among those living in rural areas. Finally, boys in rural and urban areas are 3-5 times more likely to be hospitalized for an ATV-related injury than girls. Collectively, these findings reinforce calls for specific policies and interventions aimed at preventing ATV-related injuries among rural-dwelling pediatric populations, particularly male children and adolescents.

A primary purpose of descriptive epidemiology is to identify populations at risk of disease or injury to target preventive public health measures(13). The current study provides empirical evidence to guide recommendations for preventing ATV injuries among children and adolescents by targeting public health efforts to rural dwelling youth(5, 17). Specifically, among rural youth, rates of ATV-related hospitalizations are ∼30 to 50/100,000 and among rural adolescents, the rate is 2-3 times higher than the rate of hospitalizations for motor vehicle and bicycle-related injuries, making ATV-related injuries in rural children and adolescents the most common preventable injury among adolescents in Canada. The observation that ATV-related hospitalizations are highest among adolescents supports previous work in this area, and expands it with the observation that rural-dwelling adolescents display a particularly excessive risk(19). Collectively, these data provide empirical evidence to justify the need for public health efforts aimed at reducing ATV-related injuries in rural-dwelling children and adolescents.

Geographic distributions of pediatric conditions and injuries can further refine public health efforts to prevent hospitalizations. Similar to previous studies, we found pronounced geographic differences in rural-urban inequities in hospitalizations for ATV-related injuries among children and adolescents. For example, although Ontario has one of the lowest incident rates of pediatric ATV-related hospitalizations in Canada, it was marked by the largest inequity in rural-urban differences in ATV-related hospitalizations. Conversely, the Atlantic provinces had some of the highest rates of pediatric ATV-related hospitalizations they also displayed the lowest rates of rural-urban inequities. Policies and laws for ATV-related injuries differ widely in Canada and are governed by provinces. The regional differences in the rural-urban inequities in pediatric ATV-related hospitalizations provide data to guide additional research and potential insight into future provincial policy recommendations. The current study was not designed to examine the effects of these policies on ATV-related hospitalizations, however future research could examine if differences in provincial policies contribute to some of these variations.

ATV-related injuries are overwhelmingly more common in boys than girls (4, 20, 21). These gender gaps are evident across the injury severity spectrum from fractures(20, 22) to mortality(23). American and Canadian guidelines for preventing ATV-injuries(6, 24) argue that boys have greater exposure time on ATVs and are more likely to engage in risk taking behaviours(25). We previously documented that the risks of severe injuries and death were 3 to 4-fold higher in boys and men of 16 to 20 years compared to younger and older patients, however we did not normalize the rates to the population(11). We overcome this limitation and linked population sizes to different age groups and found incidence rates are up to 8-times higher in boys than girls, particularly in rural areas. In the absence of exposure or behavioural data, the mechanism for this gender gap are unclear. These data reinforce calls for tailored public health messaging with a particular emphasis on adolescent boys and young adult men to prevent ATV-related injuries.

### Study limitations

Firstly, as with all studies that rely on large administrative databases, data are at risk of misclassification bias as only these with severe injuries and those with access to hospital were included in the analyses. Additionally, several potential confounding variables that contribute to the risk of injury severity were not available to explain the differences in ATV-related injury rates presented here. These include lifestyle-related factors like ATV-exposure time (i.e., hours of riding), use of drugs or alcohol at the time of injury, parental supervision, risky riding behaviours, purpose of ATV use (transportation vs recreational) and use of safety equipment. Data explaining the potential mechanism for the excessive rural injuries could inform public health prevention efforts. Secondly, important co-variates including driver status(20), helmet use(20, 26) and/or effect modifiers, such as type of ATV(27) were not available for all patients in this dataset. Therefore, it is unclear if differences in these variables contributed to the rural-urban differences observed here. Thirdly, these data are at risk of ascertainment bias as patients who were not seen in hospital, or died outside the hospital were not included in the analysis. This could alter the effect size and the precision of the differences, injuries rates and trends reported here. Lastly, we did not have data for the entire country, therefore the estimates of injury rates presented here could be imprecise as rural-urban inequities were not available for patients living in northern territories where ATV use could be higher.

## CONCLUSIONS

Rural dwelling children and adolescents are hospitalized for an ATV-related injury at a rate 5-8 times higher than urban dwelling children and adolescents. These geographic differences are evident for both common and rare but severe injuries. These differences are driven largely by injuries among rural boys and men. The excessive burden of ATV-related hospitalizations among rural children and adolescents reinforce calls for specific public health prevention efforts aimed at pediatric rural-dwelling ATV users, particularly adolescent boys and young adult men.

## Data Availability

All data produced in the present study are available upon reasonable request to the authors

## Abbreviations

ATV: All-terrain vehicle
ICD: international classification of diseases
DAD: discharge abstract database.

## AUTHOR CONTRIBUTIONS

Dr. McGavock had full access to all the data in the study and takes responsibility for the integrity of the data and the accuracy of the data analysis.

Concept and design: Heck, Balshaw, McGavock.

Acquisition, analysis, or interpretation of data: Sobhan, Heck, Balshaw, McGavock. Drafting of the manuscript: Heck, McGavock.

Critical revision of the manuscript for important intellectual content: All authors. Statistical analysis: Sobhan, Balshaw.

Administrative, technical, or material support: Sobhan, Heck, Balshaw, McGavock. Supervision: McGavock, Balshaw.

## DATA SHARING STATEMENT

Data can be made publicly available through an application process through the Canadian Institutes for Health Information.

## MEETING PRESENTATION

This work is not scheduled to be presented at any upcoming scientific meeting.

## ORIGINALITY OF CONTENT

All information and materials in the manuscript are original.

