## Supplemental data for "Urban-rural differences in pediatric ATV-related trauma in Canada from 2002-2019: A population-based descriptive study"

Supplemental Information

eFigure 1. Flowchart of data available and final database used for analyses

eFigure 2. Annual percentage change in hospitalizations for ATV injuries between 2003 and 2019

eFigure 3. Provincial annual incidence rates of hospitalizations for an ATV injury stratified by age group

eFigure 4. Annual percentage change by age group and geographic location

eTable 1. ICD codes used to determine injury type and calculate injury severity scores

eTable 2 : Incidence rate of ATV-related hospitalizations per 100,000 stratified by age and gender

R code for proposed analyses

eFigure 1. Flowchart of data available and final database used for analyses

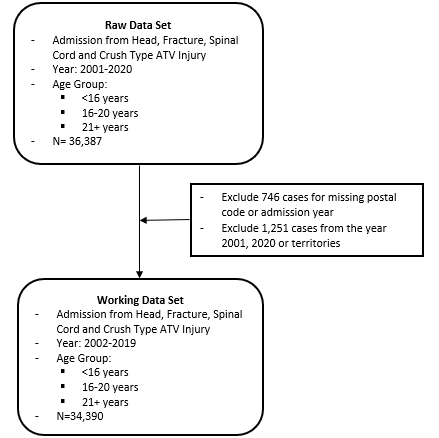

eFigure 2. Annual percentage change in hospitalizations for ATV injuries between 2003 and 2019

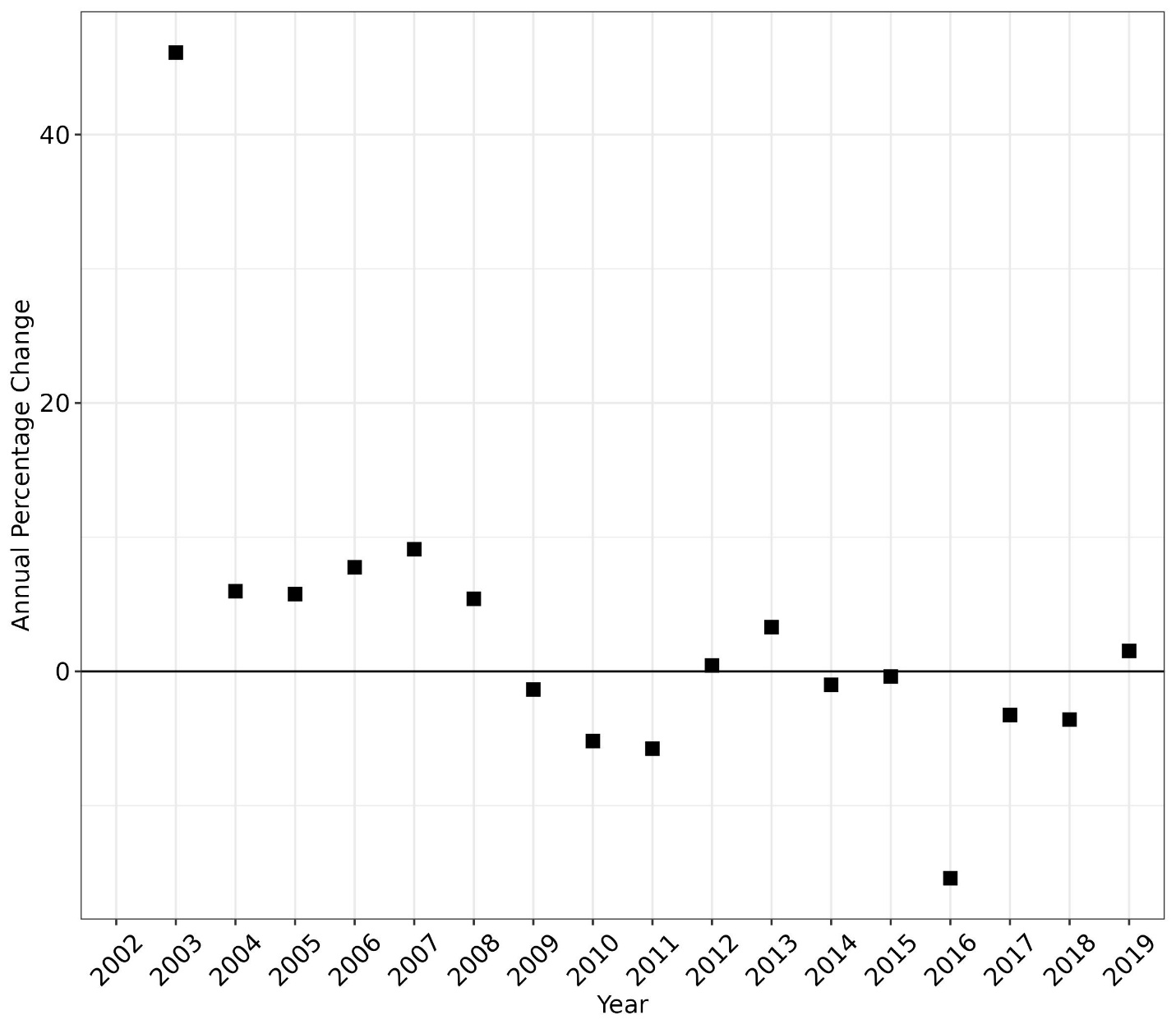

eFigure 3. Provincial annual incidence rates of hospitalizations for an ATV injury stratified by age group

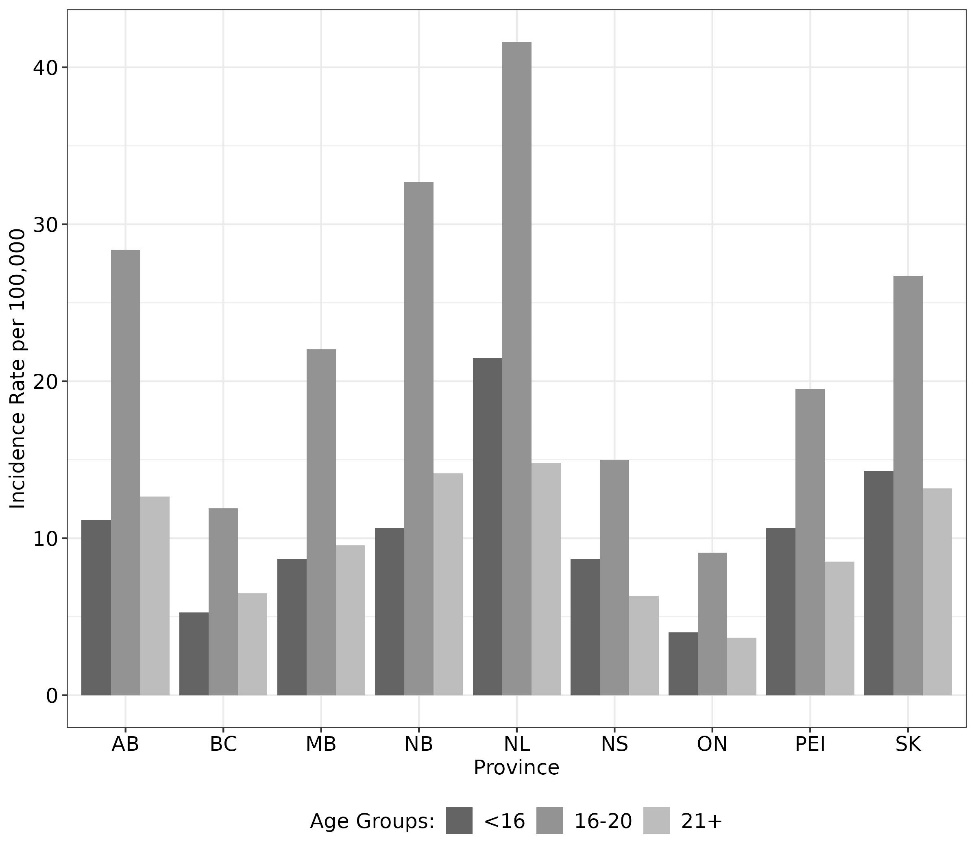

eFigure 4. Annual percentage change by age group and geographic location

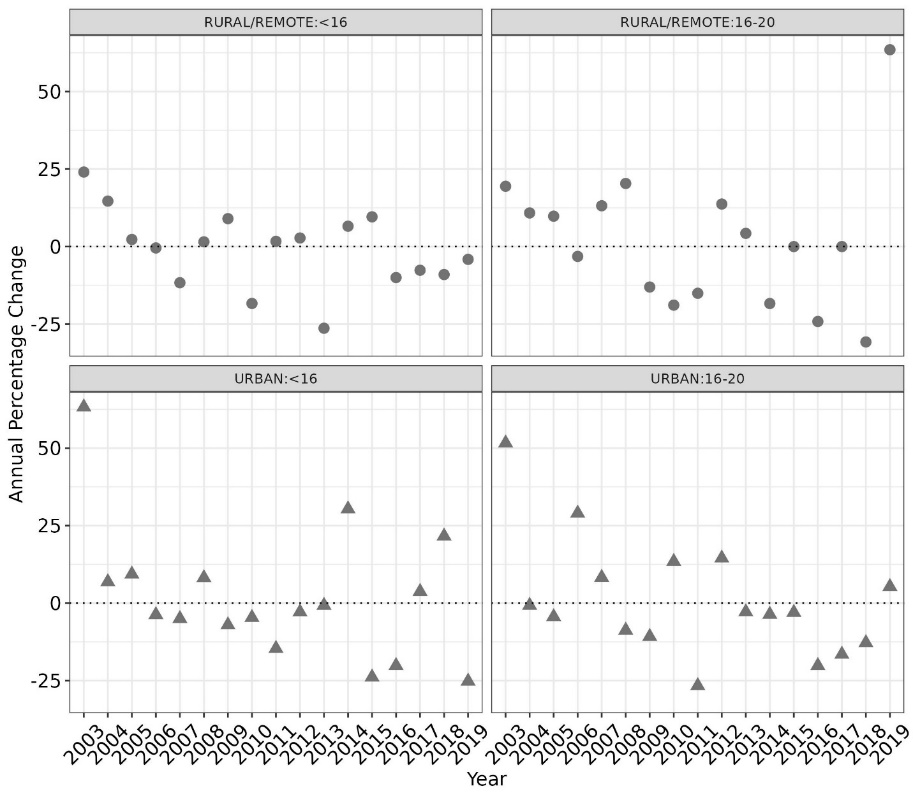

**eTable 1:** ICD codes used to determine injury type and calculate injury severity scores

| **Injury Type** | **ICD-10-CA** |
| --- | --- |
| Any ATV Injury | V86.0, V86.08-V86.98 |
| Head injury (intracranial) | S060-S069 |
| Extremity fractures | S420-S429, S520-S529, S620-S629  S720-S729, S820-S829, S920-S929  T022-T027, T10, T12 |
| Spinal cord injury (SCI with or without spinal fracture, cauda equina syndrome) | S140-S14.19, S240-S24.19, S340-S34.19, S343-S34.38 |
| Crush-type injury | S070-S079, S170-S179, S280, S380-S381, S470-S478, S570-S579, S670-S678, S770-S772, S870-S878, S970-S978, T040-T049 |

eTable 2: Incidence rate of ATV-related hospitalizations per 100,000 stratified by age and gender

| **Gender** | **Overall** | **Age Groups** | | |
| --- | --- | --- | --- | --- |
|  |  | **Children** | **Adolescent** | **Adult** |
| Male | 12 | 10 | 25 | 12 |
| Female | 2 | 3 | 5 | 2 |

R code for proposed analyses

\# Library

library(tidyverse)

library(gt)

library(gtsummary)

library(broom)

library(reshape2)

library(glmnet)

\# Read Data

combined_dad<-read_csv("~/Combined_dad_2001_2019.csv")

statcan_dat<- read.csv("~/statcan_population_data.csv",stringsAsFactors = T)

\# Preparation of Injury Data

\## Exclude missing postal code and admission year

combined_dad_v1 = combined_dad  |>

select(study_id, province_code, adm_year, age_group, gender,

urban_rural, head_injury, fracture, sci, crush, driver, passenger) |>

     	mutate_at(vars(province_code,  adm_year, age_group, gender, urban_rural,

driver, passenger), as.factor) |>

   	filter(!is.na(urban_rural))|>

   	filter(!is.na(adm_year))|>

   	filter(adm_year %in% c(2001:2020))

\## Exclude 2001, 2020 and territories

combined_dad_v2 <- combined_dad_v1|>

filter(province_code %in% c("AB", "BC", "MB", "NB", "NL", "NS", "ON", "PEI", "SK"))|>

             	filter(adm_year%in% c(2002:2019))

\## Create injury variable

combined_dad_v3 = combined_dad_v2 |>

   	column_to_rownames("study_id") |>

   	dplyr::group_by(province_code, age_group, urban_rural, gender,

                 	adm_year, driver, passenger)|>

   	dplyr::summarise(

total_head_injury = sum(head_injury),

       	total_fracture = sum(fracture),

       	total_sci = sum(sci),

       	total_crush = sum(crush)) |>

   	ungroup()

\# Prepartion of Population Data

statcan_dat_v1 = statcan_dat |>

   	dplyr::select(-reference_date) |>

   	filter(reference_year %in% c(2002:2019)) |>

   	dplyr::mutate(reference_year = as.factor(reference_year))

statcan_dat_v1$province_code <- if_else(statcan_dat_v1$province_code=="PE", "PEI",

if_else(statcan_dat_v1$province_code %in% c("YT", "NT", "NU"), "territories", statcan_dat_v1$province_code))

statcan_dat_v2 <- statcan_dat_v1|>

       	filter(province_code %in% c("AB", "BC","AB", "MB", "NB", "NS",

                                   	"NL", "SK","PEI"))

\# Merge Two Datasets (injury and population data)

combined_data = full_join(

   	combined_dad_v3, statcan_dat_v2,

   	by = c("province_code" = "province_code",

           	"age_group"    = "age_group",

           	"urban_rural"   = "urban_rural",

           	"gender"        = "gender",

           	"adm_year"      = "reference_year"))|>

dplyr::mutate(province_code =

as.factor(province_code)) |>

droplevels

\# Check for Missing

\## Note: Combined dataset has cases with no injury, e.g., BC in 2018 for Female and <16 ##group has no injury but has population data. Replace all NAs by 0

combined_data_complete_case = combined_data|>

   	mutate(across(where(is.numeric), ~replace(.,is.na(.),0)))

\# Data Management

\## Make a new column of total_any_injury to sum the different injuries

working_dat_v1 = combined_data_complete_case |>

   	mutate(total_any_injury = total_head_injury +

                                 	total_fracture + total_sci +  total_crush) |>

droplevels

\## Modify reference

working_dat_v2 = working_dat_v1 |>

   	mutate_at(vars(province_code,  age_group, gender, urban_rural),

as.factor)

   	mutate(rural_urban = if_else(urban_rural == "URBAN", 0, 1),

     	 year_f = factor(adm_year), age_grp = age_group|>

           	fct_relevel("21+"))

\# Descriptives

\## Get population size

n <- sum(working_dat_v2$total_any_injury)

n

\## Create population at risk

working_dat_v2$population1 <- working_dat_v2$population –

working_dat_v2$total_any_injury

\## Annual injury by age group

with(working_dat_v2, tapply(population1, age_grp, sum))/18

\## Annual injury by age group and year

with(working_dat_v2, tapply(population1, list(year_f, age_grp), sum))

\## Injury count and percentage by age group

with(working_dat_v2, tapply(total_any_injury, age_grp, sum))

with(working_dat_v2, tapply(total_any_injury, age_grp, sum))*100/n

\## Driver-Passenger by age group

with(working_dat_v2, tapply(driver, age_grp, sum))

with(working_dat_v2, tapply(passenger, age_grp, sum))

\## Injury count by gender

with(working_dat_v2, tapply(total_any_injury, list(gender),sum))/n

\## Injury count and percentage by age and gender

gender_by_age <- with(working_dat_v2, tapply(total_any_injury,

                                             list(gender, age_grp),sum))

gender_by_age[2,]*100/colSums(gender_by_age)

\## Injury count and percentage by age and region

region_by_age <- with(working_dat_v2, tapply(total_any_injury, list(urban_rural,

age_grp),sum))

region_by_age[1,]*100/colSums(region_by_age)

\## Injury count by region

with(working_dat_v2, tapply(total_any_injury, list(urban_rural),sum))/n

\## Injury count by age and province

province_by_age <- with(working_dat_v2, tapply(total_any_injury,

                                             	 list(province_code, age_grp),sum))

\# Annual Percentage Change (eFigure 2)

year <- 2002:2019

d <- data.frame(year = year, count = with(working_dat_v2, tapply(total_any_injury,

                                                      list(year_f),sum)))

d1 <- d|>

   	mutate(year = factor(year),

           	annual_chg = (count - lag(count))/lag(count)*100)

   	min(abs(d1$annual_chg), na.rm=TRUE);max(abs(d1$annual_chg), na.rm=TRUE)

working_dat_v2|>group_by(year_f)|>

summarize(count=sum(total_any_injury), .groups="drop")|>

     	mutate(year = factor(year),

           	annual_chg = (count - lag(count))/lag(count)*100)

pct_chg<- d1|>

ggplot(aes(x=year, y = annual_chg))+

   	theme_bw()+

   	theme(axis.text.y=element_text(size=12, face = "bold", color = "black"),

axis.text.x=element_text(angle = 45, size=12, face = "bold",

color = "black", vjust=0.5, hjust=0.5),

       	 axis.title.x=element_text(size=12, face = "bold", color = "black"),

         	axis.title.y=element_text(size=12, face = "bold", color = "black"))+

   	geom_point(shape=15, size=3, position = position_dodge(0.3)) +

   	geom_hline(yintercept=0, linetype = "solid", color = "black")+

   	xlab("Year")+ylab("Annual Percentage Change")

ggsave("pct_chg.jpg", plot = pct_chg, width =8, height = 7)

\# Annual Percentage Change by Region and Age Groups (eFigure 4)

working_dat_child_rural <- working_dat_v2|>

filter(age_grp == "<16")|>

filter(urban_rural=="RURAL/REMOTE")

working_dat_adol_rural <- working_dat_v2|>

filter(age_grp == "16-20")|>

filter(urban_rural=="RURAL/REMOTE")

working_dat_child_urban <- working_dat_v2|>

filter(age_grp == "<16")|>

filter(urban_rural=="URBAN")

working_dat_adol_urban <- working_dat_v2|>

filter(age_grp == "16-20")|>

filter(urban_rural=="URBAN")

pct_chg_child_rural <- working_dat_child_rural|>

     	group_by(year_f)|>

     	summarize(count=sum(total_any_injury), .groups="drop")|>

     	mutate(year = factor(year_f),

         	annual_chg = (count - lag(count))/lag(count)*100)

pct_chg_child_urban<- working_dat_child_urban|>

     	group_by(year_f)|>

     	summarize(count=sum(total_any_injury), .groups="drop")|>

     	mutate(year = factor(year_f),

           	annual_chg = (count - lag(count))/lag(count)*100)

pct_chg_adol_urban<- working_dat_adol_urban|>

     	group_by(year_f)|>

     	summarize(count=sum(total_any_injury), .groups="drop")|>

     	mutate(year = factor(year_f),

           	annual_chg = (count - lag(count))/lag(count)*100)

pct_chg_adol_rural<- working_dat_adol_rural|>

     	group_by(year_f)|>

     	summarize(count=sum(total_any_injury), .groups="drop")|>

     	mutate(year = factor(year_f),

           	annual_chg = (count - lag(count))/lag(count)*100)

pct_grp_tab <- data.frame(rbind(pct_chg_child_urban[-1,-c(1:2)],

pct_chg_child_rural[-1,-c(1:2)],

     	pct_chg_adol_urban[-1,-c(1:2)], pct_chg_adol_rural[-1,-c(1:2)]),

                     	`age_grp` = c(rep("<16", 34), rep("16-20", 34)),

                         	`urban_rural` = c(rep("URBAN", 17), rep("RURAL/REMOTE", 17),

                           	rep("URBAN", 17), rep("RURAL/REMOTE",17)))

pct_chg_by_grp<- pct_grp_tab|>

   	mutate(group=paste(urban_rural, age_grp, sep=":"))|>

   	ggplot(aes(x=year, y=annual_chg, group = group))+

   	facet_wrap(~group)+

   	theme_bw()+

 	theme(axis.text.y=element_text(size=12, face = "bold",

color = "black"), axis.text.x=element_text(angle = 45,

size=12, face = "bold", color = "black", vjust=0.5, hjust=0.5),

         	axis.title.x=element_text(size=12, face = "bold", color =

"black"), axis.title.y=element_text(size=12, face = "bold", color = "black"))+

geom_point(aes(shape=urban_rural), color = "grey45",size=3,

position = position_dodge(0.3)) +

   	geom_hline(yintercept=0, linetype = "dotted", color = "black")+

   	xlab("Year") + ylab("Annual Percentage Change")+

   	theme(legend.position="none")#, legend.box="horizontal")

ggsave("pct_chg_by_grp.jpg", plot = pct_chg_by_grp, width =8, height = 7)

\# Annual Incident Rate per Hundred-Thousand

\## By age-gender-region

with(working_dat_v2,tapply(total_any_injury, list(gender), sum))*100000/

with(working_dat_v2, tapply(population,list(gender), sum))

b <- working_dat_v2|>

    filter(age_grp=="16-20")

a <- working_dat_v2|>

    filter(age_grp=="<16")

c<- working_dat_v2|>

    filter(age_grp=="21+")

with(a,tapply(total_any_injury, list(gender), sum))*100000/

with(a, tapply(population,list(gender), sum))

with(b,tapply(total_any_injury, list(gender), sum))*100000/

with(b, tapply(population,list(gender), sum))

with(a,tapply(total_any_injury, list(rural_urban, gender), sum))*100000/

with(a, tapply(population,list(rural_urban, gender), sum))

with(b,tapply(total_any_injury, list(rural_urban, gender), sum))*100000/

with(b, tapply(population,list(rural_urban, gender), sum))

\## By age-gender-year

child_tab <- with(a,tapply(total_any_injury, list(gender, year_f), sum))*100000/

with(a, tapply(population,list(gender, year_f), sum))

c(min(child_tab[1,]), max(child_tab[1,])) # female

c(min(child_tab[2,]), max(child_tab[2,])) # male

adol_tab<- with(b,tapply(total_any_injury, list(gender, year_f), sum))*100000/

with(b, tapply(population,list(gender, year_f), sum))

c(min(adol_tab[1,]), max(adol_tab[1,])) # female

c(min(adol_tab[2,]), max(adol_tab[2,])) # male

\# By age and region

tapply(working_dat_v2$total_any_injury, list(working_dat_v2$urban_rural,

working_dat_v2$age_grp), sum)*100000/

tapply(working_dat_v2$population, list(working_dat_v2$urban_rural,

working_dat_v2$age_grp), sum)

tapply(working_dat_v2$total_head_injury, list(working_dat_v2$urban_rural,

working_dat_v2$age_grp), sum)*100000/

tapply(working_dat_v2$population, list(working_dat_v2$urban_rural,

working_dat_v2$age_grp), sum)

tapply(working_dat_v2$total_crush, list(working_dat_v2$urban_rural,

working_dat_v2$age_grp), sum)*100000/

tapply(working_dat_v2$population, list(working_dat_v2$urban_rural,

working_dat_v2$age_grp), sum)

tapply(working_dat_v2$total_sci, list(working_dat_v2$urban_rural,

working_dat_v2$age_grp), sum)*100000/

tapply(working_dat_v2$population, list(working_dat_v2$urban_rural,

working_dat_v2$age_grp), sum)

tapply(working_dat_v2$total_fracture, list(working_dat_v2$urban_rural,

working_dat_v2$age_grp), sum)*100000/

tapply(working_dat_v2$population, list(working_dat_v2$urban_rural,

working_dat_v2$age_grp), sum)

\## By age and year

age_by_year <- with(working_dat_v2, tapply(total_any_injury, list(age_grp, year_f),

sum))*1000/

with(working_dat_v2, tapply(population,list(age_grp, year_f), sum))

age_by_year

c(min(age_by_year[2,])*100, max(age_by_year[2,])*100)

c(min(age_by_year[3,])*100,max(age_by_year[3,])*100)

c(min(age_by_year[1,])*100,max(age_by_year[1,])*100)

\# By region and year

with(b, tapply(total_sci, list(urban_rural, year_f), sum)*100000/

tapply(population, list(urban_rural, year_f), sum))

with(b, tapply(total_sci, list(urban_rural, year_f), sum)*100000/

tapply(population, list(urban_rural, year_f), sum))

\## By province

with(working_dat_v2,tapply(total_any_injury, list(province_code), sum))

with(working_dat_v2,tapply(total_any_injury, list(province_code), sum))*100000/

with(working_dat_v2, tapply(population,list(province_code), sum))

\# By age, year and province

with(a,tapply(total_any_injury, list(province_code, year_f), sum))*100000/

with(a, tapply(population,list(province_code, year_f), sum))

with(a,tapply(total_any_injury, list(province_code), sum))*100000/

with(a, tapply(population,list(province_code), sum))

with(b,tapply(total_any_injury, list(province_code, year_f), sum))*100000/

with(b, tapply(population,list(province_code, year_f), sum))

with(b,tapply(total_any_injury, list(province_code), sum))*100000/

with(b, tapply(population,list(province_code), sum))

\## By injury type-age-region

with(working_dat_v2, tapply(total_fracture, list(age_grp,rural_urban), sum))*100000/

with(working_dat_v2, tapply(population, list(age_grp,rural_urban), sum))

with(working_dat_v2, tapply(total_head_injury, list(age_grp,rural_urban), sum))*100000/

with(working_dat_v2, tapply(population, list(age_grp,rural_urban), sum))

with(working_dat_v2, tapply(total_sci, list(age_grp,rural_urban), sum))*100000/

with(working_dat_v2, tapply(population, list(age_grp,rural_urban), sum))

with(working_dat_v2, tapply(total_crush, list(age_grp,rural_urban), sum))*100000/

with(working_dat_v2, tapply(population, list(age_grp,rural_urban), sum))

with(working_dat_v2, tapply(total_any_injury, list(age_grp,rural_urban), sum))*100000/

with(working_dat_v2, tapply(population, list(age_grp,rural_urban), sum))

\# Annual Incidence Rate by Age-Province

prov_tab <- melt(list(tapply(working_dat_v2$total_any_injury,

       	list(working_dat_v2$province_code, working_dat_v2$age_grp),

sum)*100000/

                     	tapply(working_dat_v2$population,

       	list(working_dat_v2$province_code, working_dat_v2$age_grp),

sum)))

prov_hist <- prov_tab|>

mutate(Var2=Var2|>

           	fct_relevel(c("<16", "16-20", "21+")))|>

   	ggplot()+

   	geom_bar(aes(x=Var1, y=value, fill=Var2), stat = "identity",

position="dodge")+

   	theme_bw()+

   	theme(axis.text.y=element_text(size=12, face = "bold", color = "black"),

axis.text.x=element_text(size=12, face = "bold", color = "black"),

         	axis.title.x=element_text(size=12, face = "bold", color = "black"),

         	axis.title.y=element_text(size=12, face = "bold", color = "black"),

       	legend.text=element_text(size=12, face = "bold", color = "black"),

         	legend.title = element_text(size=12, face = "bold", color = "black"))+

         	labs(x="Province", y="Incidence Rate per 100,000",

fill = "Age Groups:")+

   	theme(legend.position="bottom", legend.box="horizontal")+

   	scale_fill_manual("Age Groups:",values = c("<16"="#646464",

"16-20"="#939393",

                                               	"21+"="#BDBDBD"),

                     	limits = c("<16", "16-20", "21+"))

ggsave("prov_hist.jpg", plot = prov_hist, width =8, height = 7)

\# Figure 1

working_dat_v2_long = working_dat_v2 |>

   	pivot_longer(cols = c(total_head_injury, total_fracture, total_sci, total_crush,

total_any_injury),

       	names_to = "injury_type",

       	values_to = "cases") |>

   	mutate(injury_type = str_remove(injury_type, "total_"),

           	adm_year = as.numeric(as.character(adm_year)))

proportion_value = 1000; alpha_val = 0.5

working_dat_v2_long_per1000 = working_dat_v2_long |>

   	mutate(cases_per1000 = (cases/ population)*proportion_value)

working_dat_v2_long_per1000_var_ci <- working_dat_v2_long_per1000|>

   	mutate(# variance of incident rate (IR)

       	variance_IR = cases/(population^2),

       	# Standard error incident rate

       	SE = sqrt(variance_IR),

       	# 95% confidence interval

       	lower_bound = (cases/population -1.96*SE)*1000,

       	upper_bound = (cases/population +1.96*SE)*1000)

injury_types = c(`<16` = "<16", `16-20` = "16-20", `21+` = "21+",

`any_injury` = "Any Injury", `crush` = "Crush Injury",

`fracture` = "Fracture",`head_injury` = "Head Injury",

`sci` = "Spinal Cord Injury")

plot_gg = ggplot(working_dat_v2_long_per1000_var_ci, aes(x = adm_year,

y = cases_per1000, fill = urban_rural, color = urban_rural)) +

   	geom_line(alpha = alpha_val, linewidth =0.2) +

   	geom_point(shape=4, size=0.5, alpha=alpha_val,

position=position_dodge(width=0.2)) +

   	geom_errorbar(aes(ymin=lower_bound, ymax=upper_bound), width =0.01,

                 	alpha = alpha_val, linewidth =0.2) +

   	geom_ribbon(aes(ymin=lower_bound, ymax=upper_bound),

alpha = 0.2, color = NA) +

   	facet_grid(injury_type~age_group, scales = "free_y",

labeller = as_labeller(injury_types)) +

   	scale_color_manual(values = c("RURAL/REMOTE"="#939393",

"URBAN" = "#505050"), limits = c("RURAL/REMOTE", "URBAN"))+

   	scale_fill_manual(values = c("RURAL/REMOTE"="#939393", "URBAN"="#505050"),

                      limits = c("RURAL/REMOTE", "URBAN"))+

   	theme_bw() +

   	theme(axis.text.x = element_text(angle =45, hjust = 1),

         	plot.title=element_text(hjust=0.5), strip.text=element_text(size = 8)) +

   	ggtitle("Injury Proportions for \n NS, ON, NL, NB, SK, BC, AB, PEI, MB Provinces")+

   	labs(x= "Admission Year", y= "Cases per 1000", fill="Urban/Rural",

color = "Urban/Rural")

ggsave("population_adj_injury_rate_9prov.jpg", plot = plot_gg, width =8, height = 7)

\# Figure 2 (incidence rate ratio by province)

AB<- working_dat_v2|>filter(province_code=="AB")

BC<- working_dat_v2|>filter(province_code=="BC")

MB<- working_dat_v2|>filter(province_code=="MB")

NB<- working_dat_v2|>filter(province_code=="NB")

NS<- working_dat_v2|>filter(province_code=="NS")

NL<- working_dat_v2|>filter(province_code=="NL")

SK<- working_dat_v2|>filter(province_code=="SK")

PEI<- working_dat_v2|>filter(province_code=="PEI")

ON<- working_dat_v2|>filter(province_code=="ON")

mAB1<- glm(total_any_injury ~ rural_urban  + year_f, subset= (age_grp=="<16"),

         	family = poisson, data = AB, offset = log(population))

mAB2<- glm(total_any_injury ~ rural_urban  + year_f, subset= (age_grp=="16-20"),

         	family = poisson, data = AB, offset = log(population))

mBC1<- glm(total_any_injury ~ rural_urban  + year_f, subset= (age_grp=="<16"),

         	family = poisson, data = BC, offset = log(population))

mBC2<- glm(total_any_injury ~ rural_urban  + year_f, subset= (age_grp=="16-20"),

         	family = poisson, data = BC, offset = log(population))

mMB1<- glm(total_any_injury ~ rural_urban  + year_f, subset= (age_grp=="<16"),

         	family = poisson, data = MB, offset = log(population))

mMB2<- glm(total_any_injury ~ rural_urban  + year_f, subset= (age_grp=="16-20"),

         	family = poisson, data = MB, offset = log(population))

mNB1<- glm(total_any_injury ~ rural_urban  + year_f, subset= (age_grp=="<16"),

         	family = poisson, data = NB, offset = log(population))

mNB2<- glm(total_any_injury ~ rural_urban  + year_f, subset= (age_grp=="16-20"),

         	family = poisson, data = NB, offset = log(population))

mNL1<- glm(total_any_injury ~ rural_urban  + year_f, subset= (age_grp=="<16"),

         	family = poisson, data = NL, offset = log(population))

mNL2<- glm(total_any_injury ~ rural_urban  + year_f, subset= (age_grp=="16-20"),

           	family = poisson, data = NL, offset = log(population))

mNS1<- glm(total_any_injury ~ rural_urban  + year_f, subset= (age_grp=="<16"),

         	family = poisson, data = NS, offset = log(population))

mNS2<- glm(total_any_injury ~ rural_urban  + year_f, subset= (age_grp=="16-20"),

         	family = poisson, data = NS, offset = log(population))

mON1<- glm(total_any_injury ~ rural_urban  + year_f, subset= (age_grp=="<16"),

         	family = poisson, data = ON, offset = log(population))

mON2<- glm(total_any_injury ~ rural_urban  + year_f, subset= (age_grp=="16-20"),

         	family = poisson, data = ON, offset = log(population))

mSK1<- glm(total_any_injury ~ rural_urban  + year_f, subset= (age_grp=="<16"),

         	family = poisson, data = SK, offset = log(population))

mSK2<- glm(total_any_injury ~ rural_urban  + year_f, subset= (age_grp=="16-20"),

         	family = poisson, data = SK, offset = log(population))

mPEI1<- glm(total_any_injury ~ rural_urban  + year_f, subset= (age_grp=="<16"),

         	family = poisson, data = PEI, offset = log(population))

mPEI2<- glm(total_any_injury ~ rural_urban  + year_f, subset= (age_grp=="16-20"),

         	family = poisson, data = PEI, offset = log(population))

NL_res1 <- mNL1|>

tidy(Std.Error=TRUE)|>

filter(term=="rural_urban")|>

select(term, estimate, se = std.error)|>

mutate(or = exp(estimate), conf.low = exp(estimate - 1.96*se),

     	conf.high = exp(estimate + 1.96*se))

NL_res2 <- mNL2|>

tidy(Std.Error=TRUE)|>

filter(term=="rural_urban")|>

select(term, estimate, se = std.error)|>

mutate(or = exp(estimate), conf.low = exp(estimate - 1.96*se),

     	conf.high = exp(estimate + 1.96*se))

NB_res1 <- mNB1|>

tidy(Std.Error=TRUE)|>

filter(term=="rural_urban")|>

select(term, estimate, se = std.error)|>

mutate(or = exp(estimate), conf.low = exp(estimate - 1.96*se),

     	conf.high = exp(estimate + 1.96*se))

NB_res2 <- mNB2|>

tidy(Std.Error=TRUE)|>

filter(term=="rural_urban")|>

select(term, estimate, se = std.error)|>

mutate(or = exp(estimate), conf.low = exp(estimate - 1.96*se),

     	conf.high = exp(estimate + 1.96*se))

MB_res1 <- mMB1|>

tidy(Std.Error=TRUE)|>

filter(term=="rural_urban")|>

select(term, estimate,se = std.error)|>

mutate(or = exp(estimate), conf.low = exp(estimate - 1.96*se),

     	conf.high = exp(estimate + 1.96*se))

MB_res2 <- mMB2|>

tidy(Std.Error=TRUE)|>

filter(term=="rural_urban")|>

select(term, estimate,se = std.error)|>

mutate(or = exp(estimate), conf.low = exp(estimate - 1.96*se),

     	conf.high = exp(estimate + 1.96*se))

NS_res1 <- mNS1|>

tidy(Std.Error=TRUE)|>

filter(term=="rural_urban")|>

select(term, estimate, se = std.error)|>

mutate(or = exp(estimate), conf.low = exp(estimate - 1.96*se),

     	conf.high = exp(estimate + 1.96*se))

NS_res2 <- mNS2|>

tidy(Std.Error=TRUE)|>

filter(term=="rural_urban")|>

select(term, estimate, se = std.error)|>

mutate(or = exp(estimate), conf.low = exp(estimate - 1.96*se),

     	conf.high = exp(estimate + 1.96*se))

ON_res1 <- mON1|>

tidy(Std.Error=TRUE)|>

filter(term=="rural_urban")|>

select(term, estimate, se = std.error)|>

mutate(or = exp(estimate), conf.low = exp(estimate - 1.96*se),

     	conf.high = exp(estimate + 1.96*se))

ON_res2 <- mON2|>

tidy(Std.Error=TRUE)|>

filter(term=="rural_urban")|>

select(term, estimate, se = std.error)|>

mutate(or = exp(estimate), conf.low = exp(estimate - 1.96*se),

     	conf.high = exp(estimate + 1.96*se))

SK_res1 <- mSK1|>

tidy(Std.Error=TRUE)|>

filter(term=="rural_urban")|>

select(term, estimate, se = std.error)|>

mutate(or = exp(estimate), conf.low = exp(estimate - 1.96*se),

     	conf.high = exp(estimate + 1.96*se))

SK_res2 <- mSK2|>

tidy(Std.Error=TRUE)|>

filter(term=="rural_urban")|>

select(term, estimate, se = std.error)|>

mutate(or = exp(estimate), conf.low = exp(estimate - 1.96*se),

     	conf.high = exp(estimate + 1.96*se))

PEI_res1 <- mPEI1|>

tidy(Std.Error=TRUE)|>

filter(term=="rural_urban")|>

select(term, estimate,se = std.error)|>

mutate(or = exp(estimate), conf.low = exp(estimate - 1.96*se),

     	conf.high = exp(estimate + 1.96*se))

PEI_res2 <- mPEI2|>

tidy(Std.Error=TRUE)|>

filter(term=="rural_urban")|>

select(term, estimate,se = std.error)|>

mutate(or = exp(estimate), conf.low = exp(estimate - 1.96*se),

     	conf.high = exp(estimate + 1.96*se))

AB_res1 <- mAB1|>

tidy(Std.Error=TRUE)|>

filter(term=="rural_urban")|>

select(term, estimate, se = std.error)|>

mutate(or = exp(estimate), conf.low = exp(estimate - 1.96*se),

     	conf.high = exp(estimate + 1.96*se))

AB_res2 <- mAB2|>

tidy(Std.Error=TRUE)|>

filter(term=="rural_urban")|>

select(term, estimate, se = std.error)|>

mutate(or = exp(estimate), conf.low = exp(estimate - 1.96*se),

     	conf.high = exp(estimate + 1.96*se))

BC_res1 <- mBC1|>

tidy(Std.Error=TRUE)|>

filter(term=="rural_urban")|>

select(term, estimate, se = std.error)|>

mutate(or = exp(estimate), conf.low = exp(estimate - 1.96*se),

     	conf.high = exp(estimate + 1.96*se))

BC_res2 <- mBC2|>

tidy(Std.Error=TRUE)|>

filter(term=="rural_urban")|>

select(term, estimate, se = std.error)|>

mutate(or = exp(estimate), conf.low = exp(estimate - 1.96*se),

     	conf.high = exp(estimate + 1.96*se))

prov_tab <- data.frame(province = c("AB", "AB", "BC", "BC", "MB","MB","NB","NB","NL",

"NL", "NS","NS", "ON", "ON", "PEI", "PEI", "SK","SK"),

                     	 age_grp =rep(c("<16", "16-20"), 9),

rbind(AB_res1,AB_res2, BC_res1, BC_res2, MB_res1,MB_res2,

NB_res1,NB_res2, NL_res1,  NL_res2, NS_res1, NS_res2,

ON_res1, ON_res2, PEI_res1, PEI_res2, SK_res1, SK_res2))

\## compare rural irr of age groups by province with urban counter part

prov_plot <- prov_tab[-c(3:5)]|>

   	mutate(province = factor(province, levels = c("ON","BC", "AB",

                                                  "MB", "SK", "NS", "NB", "NL", "PEI")))|>

   	ggplot(aes(y=fct_rev(province), group = age_grp))+

   	theme_bw()+

   	theme(axis.text.y=element_text(size=12, face = "bold", color = "black"),

         	axis.text.x=element_text(size=12, face = "bold", color = "black"),

         	axis.title.x=element_text(size=12, face = "bold", color = "black"),

         	axis.title.y=element_text(size=12, face = "bold", color = "black"))+

geom_point(aes(x=or, shape = age_grp, color=age_grp), size=3, position =

position_dodge(0.3)) + geom_errorbar(aes(xmin=conf.low, xmax=conf.high, linetype = age_grp, color=age_grp), width=0, lwd=1,

                 	position = position_dodge(0.3))+

   	geom_vline(xintercept=1, linetype = "dashed", color = "black")+

   	xlab("Incidence Rate Ratio (IRR)") + ylab("")+

 	scale_color_manual("Age Groups:",values = c("<16"="#999999",

"16-20"="#595959"), limits = c("<16", "16-20"))+

   	scale_shape_manual("Age Groups:",values = c("<16"=19, "16-20"=15),

                     	limits = c("<16", "16-20"))+

scale_linetype_manual("Age Groups:",values = c("<16"="solid",

"16-20"="longdash"), limits = c("<16", "16-20"))+

   	theme(legend.position="bottom", legend.box="horizontal")+xlim(0,NA)

ggsave("prov_plot.jpg", plot = prov_plot, width =10, height = 7)

\# Injury Rate by Province (eFigure 3)

\## Incidence rate per province

prov_tab2 <- melt(list(tapply(working_dat_v2$total_any_injury,

     	 list(working_dat_v2$province_code, working_dat_v2$age_grp),

sum)*100000/

                     	tapply(working_dat_v2$population,

       	list(working_dat_v2$province_code, working_dat_v2$age_grp), sum)))

prov_hist <- prov_tab2|>

mutate(Var2=Var2|>

           	fct_relevel(c("<16", "16-20", "21+")))|>

   	ggplot()+

   	geom_bar(aes(x=Var1, y=value, fill=Var2),

stat = "identity", position="dodge")+

   	theme_bw()+

   	theme(axis.text.y=element_text(size=12, face = "bold", color = "black"),

         	axis.text.x=element_text(size=12, face = "bold", color = "black"),

         	axis.title.x=element_text(size=12, face = "bold", color = "black"),

         	axis.title.y=element_text(size=12, face = "bold", color = "black"),

       	legend.text=element_text(size=12, face = "bold", color = "black"),

         	legend.title = element_text(size=12, face = "bold", color = "black"))+

         	labs(x="Province", y="Total Injury", fill = "Age Groups:")+

   	theme(legend.position="bottom", legend.box="horizontal")

\# Model Overall

\# Model1: count~rural_urban+year by age group (subset)

m1A<- glm(total_any_injury ~ rural_urban+ year_f , subset= (age_group=="<16"),

         	family = poisson, data = working_dat_v2, offset = log(population))

m1A|>

tidy(exp = TRUE, conf.int = TRUE) |>

select(term, or = estimate, conf.low, conf.high)|>

   	filter(term == "rural_urban")

m1B <- glm(total_any_injury ~ rural_urban + year_f, subset= (age_group == "16-20"),

         	family = poisson, data = working_dat_v2, offset = log(population),

           	start = rep(0, 19))

m1B|>

   	tidy(exp = TRUE, conf.int = TRUE) |>

select(term, or = estimate, conf.low, conf.high)%>%

   	filter(term == "rural_urban")

m1C <- glm(total_any_injury ~ rural_urban + year_f, subset= (age_group == "21+"),

         	family = poisson, data = working_dat_v2, offset = log(population))

m1C|>

   	tidy(exp = TRUE, conf.int = TRUE) |>

select(term, or = estimate, conf.low, conf.high)|>

   	filter(term == "rural_urban")

\# Model by gender

male_child<- glm(total_any_injury ~ gender + year_f, subset = (age_grp == "<16"),

         	family = poisson, data = working_dat_v2, offset = log(population))

male_adol<- glm(total_any_injury ~ gender + year_f, subset= (age_grp == "16-20"),

         	family = poisson, data = working_dat_v2, offset = log(population))

male_child|>

   	tidy(exp = TRUE, conf.int = TRUE) |>

select(term, or = estimate, conf.low, conf.high)%>%

   	filter(term == "genderM")

male_adol|>

tidy(exp = TRUE, conf.int = TRUE) |>

select(term, or = estimate, conf.low, conf.high)%>%

   	filter(term == "genderM")

male_child_region<- glm(total_any_injury ~ gender:rural_urban + year_f,

subset = (age_grp == "<16"), family = poisson, data = working_dat_v2, offset = log(population))

male_adol_region<- glm(total_any_injury ~ gender:rural_urban + year_f,

subset= (age_grp == "16-20"), family = poisson, data = working_dat_v2, offset = log(population))

male_child_region|>

   	tidy(exp = TRUE, conf.int = TRUE) |>

select(term, or = estimate, conf.low, conf.high)

male_adol_region|>

tidy(exp = TRUE, conf.int = TRUE) |>

select(term, or = estimate, conf.low, conf.high)

\# Model by Injury

\## Model1_crush: count~rural_urban+year by age group (subset) for crush

m1crush <- glm(total_crush ~ rural_urban + year_f, subset= (age_group == "<16"),

             	family = poisson, data = working_dat_v2, offset = log(population))

m1crush|>

tidy(Std.Error=TRUE)|>

filter(term %in% c("rural_urban"))|>

select(term, estimate,se = std.error)|>

mutate(or = exp(estimate), conf.low = exp(estimate - 1.96*se),

     	conf.high = exp(estimate + 1.96*se))

m2crush <- glm(total_crush ~ rural_urban + year_f, subset= (age_group == "16-20"),

             	family = poisson, data = working_dat_v2, offset = log(population))

m2crush|>

tidy(Std.Error=TRUE)|>

filter(term %in% c("rural_urban"))|>

select(term, estimate,se = std.error)|>

mutate(or = exp(estimate), conf.low = exp(estimate - 1.96*se),

     	conf.high = exp(estimate + 1.96*se))

\## Model1_head: count~rural_urban+year by age group (subset) for head

m1head <- glm(total_head_injury ~ rural_urban + year_f, subset= (age_group == "<16"),

             	family = poisson, data = working_dat_v2, offset = log(population))

m1head|>

tidy(Std.Error=TRUE)|>

filter(term %in% c("rural_urban"))|>

select(term, estimate,se = std.error)|>

mutate(or = exp(estimate), conf.low = exp(estimate - 1.96*se),

     	conf.high = exp(estimate + 1.96*se))

m2head <- glm(total_head_injury ~ rural_urban + year_f, subset= (age_group == "16-20"),

             	family = poisson, data = working_dat_v2, offset = log(population))

m2head|>

tidy(Std.Error=TRUE)|>

filter(term %in% c("rural_urban"))|>

select(term, estimate,se = std.error)|>

mutate(or = exp(estimate), conf.low = exp(estimate - 1.96*se),

     	conf.high = exp(estimate + 1.96*se))

\## Model1_fracture: count~rural_urban+year by age group (subset) for fracture

m1fracture <- glm(total_fracture ~ rural_urban + year_f, subset= (age_group == "<16"),

             	family = poisson, data = working_dat_v2, offset = log(population))

m1fracture|>tidy(Std.Error=TRUE)|>

filter(term %in% c("rural_urban"))|>

select(term, estimate,se = std.error)|>

mutate(or = exp(estimate), conf.low = exp(estimate - 1.96*se),

     	conf.high = exp(estimate + 1.96*se))

m2fracture <- glm(total_fracture ~ rural_urban + year_f, subset= (age_group == "16-20"),

             	family = poisson, data = working_dat_v2, offset = log(population))

m2fracture|>

tidy(Std.Error=TRUE)|>

filter(term %in% c("rural_urban"))|>

select(term, estimate,se = std.error)|>

mutate(or = exp(estimate), conf.low = exp(estimate - 1.96*se),

     	conf.high = exp(estimate + 1.96*se))

\## Model1_sci: count~rural_urban+year by age group (subset) for sci

m1sci <- glm(total_sci ~ rural_urban + year_f, subset= (age_group == "<16"),

             	family = poisson, data = working_dat_v2, offset = log(population))

m1sci|>

tidy(Std.Error=TRUE)|>

filter(term %in% c("rural_urban"))|>

select(term, estimate,se = std.error)|>

mutate(or = exp(estimate), conf.low = exp(estimate - 1.96*se),

     	conf.high = exp(estimate + 1.96*se))

m2sci <- glm(total_sci ~ rural_urban + year_f, subset= (age_group == "16-20"),

             	family = poisson, data = working_dat_v2, offset = log(population))

m2sci|>

tidy(Std.Error=TRUE)|>

filter(term %in% c("rural_urban"))|>

select(term, estimate,se = std.error)|>

mutate(or = exp(estimate), conf.low = exp(estimate - 1.96*se),

     	conf.high = exp(estimate + 1.96*se))
